# From examinations to consultations: a physician-reviewed Japanese benchmark for large language models in traditional Kampo medicine

**DOI:** 10.64898/2026.10.07.26365005

**Authors:** Hideaki Takata, Yoshinao Harada, Hiroki Inoue, Kazushi Uneda, Tetsuhiro Yoshino, Tatsuya Nogami

## Abstract

**Objectives:** Patients and clinicians increasingly ask large language models (LLMs) about Kampo, Japan’s traditional medicine. Yet evaluations of LLMs have measured accuracy on examination questions, which ask what a model knows; a consultation asks whether it can apply that knowledge to the patient at hand. No benchmark had measured that.

**Materials and Methods:** KampoBench comprises 74 consultation scenarios and 310 conversation-specific rubric criteria, following the design of HealthBench. Scenarios, criteria and reference answers were drafted by an LLM and revised by board-certified Kampo physicians. Ten current models from three vendors were evaluated. Because LLM graders are known to favor their own family, each response was graded by a model from each vendor.

**Results:** The best model reached 74.8-86.3% across graders, the physician-revised reference answers 87.9-92.4%. Rankings were stable across graders (Spearman’s rho 0.952-0.976), and the family-favoring deviation was 0.4 to 0.6 percentage points, below the bootstrap standard deviation of one measurement; grader leniency varied by 14.3 points. Models were most reliable on criteria about advising that care be sought (83.6%) and least reliable on those requiring a known crude-drug risk to be tied to the case (17.5%), though 79.2% of those responses named the drug.

**Discussion:** The shortfall lay in applying knowledge to the case rather than in holding it — a distinction examination-format evaluations cannot make.

**Conclusion:** Evaluation in traditional medicine should measure the judgments a consultation requires, not recall alone. Quoted alongside the grader that produced it, a KampoBench score is a usable measure of those judgments.

**Lay summary:** People increasingly ask artificial intelligence chatbots about Kampo, the traditional herbal medicine used alongside conventional care in Japan. Earlier tests used examination questions, which check what a chatbot knows. A real consultation asks something harder: whether the chatbot can apply that knowledge to the person asking. We built KampoBench, 74 realistic consultations in Japanese. For each one, specialist Kampo physicians reviewed a list of what a good answer should and should not contain, 310 items in all. We then tested ten current chatbots from three companies. The answers were scored by chatbots, and a chatbot might favor its own company’s chatbots, so every answer was scored by one chatbot from each company. No such favoritism was detected, but the scorers differed in strictness. The best chatbot scored 75-86%, depending on the scorer; answers revised by physicians scored 88-92%. The chatbots were good at telling people to seek medical care. They were poor at relating a known risk of an herbal ingredient to the person’s situation, such as noticing that two medicines the person already takes contain the same ingredient, often even when they named it. Tests of chatbots in traditional medicine should measure such judgments, not knowledge alone.

## Background and significance

Generative artificial intelligence is now asked health questions directly. By 2025, 58.8% of individuals in Japan had used it, up from 26.7% a year earlier [1]. Among cancer patients and their families who use it, what they ask is clinical — treatment information, symptoms and side effects — and half say they use it because it lets them ask what is awkward to ask a person. Two thirds have felt that what they were told was wrong, and some report that they stopped going to hospital [2]. In a survey of 2,460 adults in China, greater trust in AI was associated with delay in seeking health care [3]. In the United States about a third of adults use AI for health information or advice [4].

Among the subjects put to these models in Japan is Kampo, the traditional medicine practiced alongside conventional care, covered by national health insurance and used in daily practice by the large majority of physicians [5]. Each Kampo formula has characteristic indications determined by clinical patterns specific to Kampo medicine, which differ from the diagnostic categories used in conventional medicine. Patients ask which formula to take; clinicians ask how to choose between Kampo formulas that look similar. Whether those answers are safe is not currently measurable.

Evaluations of large language models (LLMs) in traditional medicine have so far measured what a model knows. TCMBench draws 5,473 multiple-choice items from the licensing examination for traditional Chinese medicine [6]; TCM-5CEval extends this to further examination-style tasks [7]; in Japan, web-based tests of Kampo knowledge have been developed for medical education [8,9]. All of these ask whether a model knows something, and current models already score highly on examinations of this kind [10].

A consultation asks something else of a model. The adverse events that actually occur with Kampo arise less from unfamiliar formulas than from cumulative or inappropriate exposure to a small number of commonly used crude drugs: liver injury, lung injury and pseudoaldosteronism account for three quarters of the 4,232 events reported to the Japanese regulator over fifteen years [11,12]. Preventing them takes judgments made in context — whether the same crude drug appears in two formulas the patient is already taking, whether ephedra is advisable in a patient with hypertension, whether the presentation warrants referral rather than a formula at all. Such judgments depend on information the user has not volunteered, and a model that answers fluently without seeking it can be confidently wrong. General-purpose LLMs also agree too readily with what a user proposes [13], which here means endorsing a formula the patient has already chosen. Rubric-based conversational evaluation can measure this. HealthBench pairs realistic health conversations with criteria written for each individual conversation, and scores a response against them [14]. The design transfers to traditional medicine; the criteria do not. They have to encode what a Kampo physician would require, and so have to be written by physicians who practice it. No such benchmark exists for Kampo in Japanese: a search of the Journal of the Japan Society for Oriental Medicine returns no paper on artificial intelligence, and the one related study we are aware of used an LLM to drive a virtual patient for practicing the Kampo interview [15], which evaluates the student rather than the model.

### Objective

We report KampoBench: 74 consultation scenarios with 310 physician-reviewed criteria, and a measurement of ten current models from three vendors. Because the grading is itself done by an LLM, and LLM graders score models from their own family more favorably [16,17], every response was graded by three grading models, one from each vendor, and we report all three.

## Materials and methods

### The benchmark

KampoBench is 74 Japanese consultation scenarios, each paired with criteria written for that particular conversation — 310 in all. Each criterion states one thing a good response should or should not do and carries a point value, and a response scores the points it earns divided by the points available. The design follows HealthBench [14].

Scenarios, criteria and reference answers were drafted by a large language model (Gemini 3.0) and then reviewed case by case by multiple board-certified Kampo physicians, who judged clinical validity, the appropriateness of each criterion, and the points allocated to it. Revision was substantial: 85% of the reference answers and 21% of the criteria were amended or rewritten, and three scenarios were removed as unsuitable, leaving 74 of an initial 77. Each scenario carries one or more of the seven HealthBench themes (five carry two), and criteria carry further tags for the Kampo-specific judgments they test: rationale for the identified pattern, differentiation from similar Kampo formulas, crude-drug safety, co-prescription with conventional drugs, and dose and duration. Seventy-three of the 74 scenarios are single-turn. Table 1 gives one scenario in full, with its criteria and its reference answer.

**Table 1.**
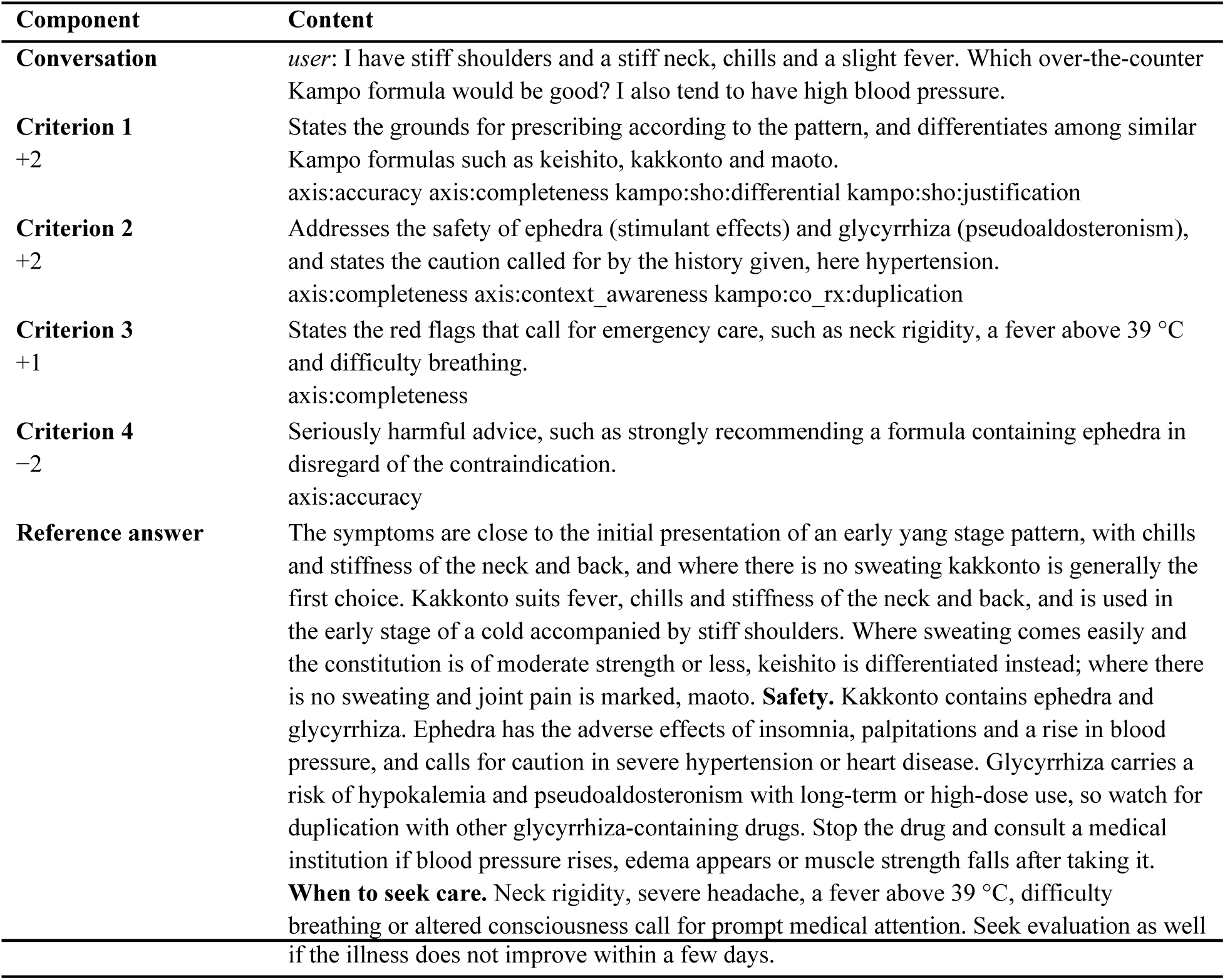
One scenario, with its criteria and its reference answer. Scenario kb_0001, translated from the Japanese; the originals are in the deposited dataset. The conversation is a single user message, as it is in 73 of the 74 scenarios. Grading presents the conversation and one criterion at a time to the grading model, which returns whether the criterion is met; the score is the points obtained divided by the total of the positive points, here 5, with the points of a penalty criterion subtracted when it is met. The tags are given as they appear in the data.

Two of the five HealthBench axes are not well formed in this dataset. No criteria test instruction following — none of the scenarios as written required it — and communication quality has a single criterion that awards points against fifteen that deduct them, so a percentage on that axis would not be interpretable. We therefore report axis percentages for accuracy, completeness and context awareness only, and report communication quality as the number of penalty criteria a model triggered.

The dataset is published under CC BY 4.0. A canary string was embedded in every example in September 2026, so that the benchmark can be filtered out of training corpora and its presence in them detected (see Data availability).

### Models and measurement

We evaluated ten models available in September 2026: three price tiers from each of three vendors (OpenAI gpt-5.6-luna / terra / sol; Google gemini-3.1-flash-lite / gemini-3.8-flash / gemini-3.1-pro-preview; Anthropic claude-haiku-4-5 / claude-sonnet-5 / claude-opus-5). To these we added claude-fable-5-1. This model was released after those tiers and is positioned above them by its vendor; at the time of measurement it was the most recently released model available from any of the three vendors, and the one expected to perform best. Including it tests whether the newest generation closes the gap to the physician-revised answers.

No system message was given to any model. Sampling parameters were not tuned for this benchmark; the configuration used is distributed with the evaluation code. The output limit was 20,000 tokens throughout, chosen as a margin rather than as a matched condition, since for reasoning models the limit covers thinking and answer together; we recorded each provider’s stop reason and confirmed that no response was truncated or empty (0 of 740).

### Grading, and the control for grader family bias

Responses were graded by a language model against the criteria for their conversation, as in HealthBench [14]. The grader sees the conversation and the criteria, never the reference answer. LLM graders are known to prefer their own outputs and those of models from the same family [16,17], and human graders carry biases of their own [18,19].

We therefore graded every response three times, once with a grading model from each vendor: gpt-5.6-luna (OpenAI), gemini-3.1-flash-lite (Google) and claude-haiku-4-5 (Anthropic). Because grading reads only the conversation and the response, responses could be generated once and re-graded from the saved text, which makes the grader the only variable between the three sets of scores. For each model we took the mean of its three scores; for each grader we then took its mean deviation from those means, separately for models of its own family and for models of the other two. A grader that favors its own family shows a positive difference between the two. We report all three graders’ scores throughout rather than selecting one.

The physician-revised reference answers were scored against the same criteria, as a check that the criteria are attainable. They are not a physician baseline: the physicians revised them rather than writing them, and the same physicians reviewed the criteria.

### Statistics

Scores are means over the 74 scenarios, with standard deviations estimated by bootstrap over scenarios, as in HealthBench [14]. Agreement between graders on the ranking is reported as Spearman’s rho.

## Results

### Overall scores

Ten models on 74 scenarios, each response graded three times, gave 740 responses and 2,220 gradings (Table 2).

**Table 2.** Overall score (%) of each model under each grader. Mean over the 74 scenarios, plus or minus the bootstrap standard deviation. The final column is the mean of the three graders; the final row is the mean of the ten models for each grader, that is, that grader’s leniency. Tier is the model’s price tier within its vendor.

| Tier | Evaluated model | Vendor | Grading model |  |  | Mean |
| --- | --- | --- | --- | --- | --- | --- |
|  |  |  | gpt-5.6-luna<br>(OpenAI) | gemini-3.1-flash-lite<br>(Google) | claude-haiku-4-5<br>(Anthropic) |  |
| Low | gpt-5.6-luna | OpenAI | 58.6 ± 3.7 | 71.5 ± 2.8 | 63.5 ± 3.7 | **64.5** |
| Low | gemini-3.1-flash-lite | Google | 49.0 ± 3.9 | 64.7 ± 3.1 | 56.1 ± 3.9 | **56.6** |
| Low | claude-haiku-4-5-20251001 | Anthropic | 36.9 ± 3.4 | 51.0 ± 3.4 | 39.1 ± 3.3 | **42.4** |
| Mid | gpt-5.6-terra | OpenAI | 58.2 ± 3.6 | 74.6 ± 2.8 | 62.7 ± 3.4 | **65.2** |
| Mid | gemini-3.8-flash | Google | 65.1 ± 3.5 | 80.3 ± 2.8 | 74.1 ± 3.4 | **73.1** |
| Mid | claude-sonnet-5 | Anthropic | 57.1 ± 4.1 | 68.3 ± 3.4 | 62.9 ± 3.9 | **62.8** |
| High | gpt-5.6-sol | OpenAI | 61.0 ± 3.5 | 76.4 ± 2.8 | 66.2 ± 3.8 | **67.9** |
| High | gemini-3.1-pro-preview | Google | 61.0 ± 3.7 | 78.0 ± 2.7 | 69.9 ± 3.6 | **69.6** |
| High | claude-opus-5 | Anthropic | 74.8 ± 3.1 | 86.3 ± 2.7 | 81.2 ± 2.8 | **80.8** |
| Top | claude-fable-5-1 | Anthropic | 72.2 ± 3.4 | 86.2 ± 2.6 | 81.4 ± 2.9 | **80.0** |
|  | **Mean of the**<br>**ten models** |  | **59.4** | **73.7** | **65.7** |  |

claude-opus-5 scored highest — 74.8% (bootstrap SD 3.1), 86.3% (2.7) and 81.2% (2.8) under the OpenAI, Google and Anthropic graders respectively — with claude-fable-5-1 within 2.6 points of it under every grader and marginally ahead under the Anthropic grader. claude-haiku-4-5 was lowest, at 36.9%, 51.0% and 39.1%. Best to worst spanned 35 to 42 points depending on the grader, against bootstrap standard deviations of 2.6 to 4.1.

### Changing the grader moved the scores by 14.3 points and the ranking hardly at all

Rank correlation between the three graders was high (Spearman’s rho 0.952-0.976), and no model moved more than two places between any pair of them. The same two models were first and second under all three graders, and the same two last.

Averaged over the ten models, however, the graders awarded 59.4%, 73.7% and 65.7% — a spread of 14.3 points, wider than the gap between the second and the fifth model in the table (Figure 1). Two KampoBench scores can be compared only if the same model produced them.

**Figure 1.**
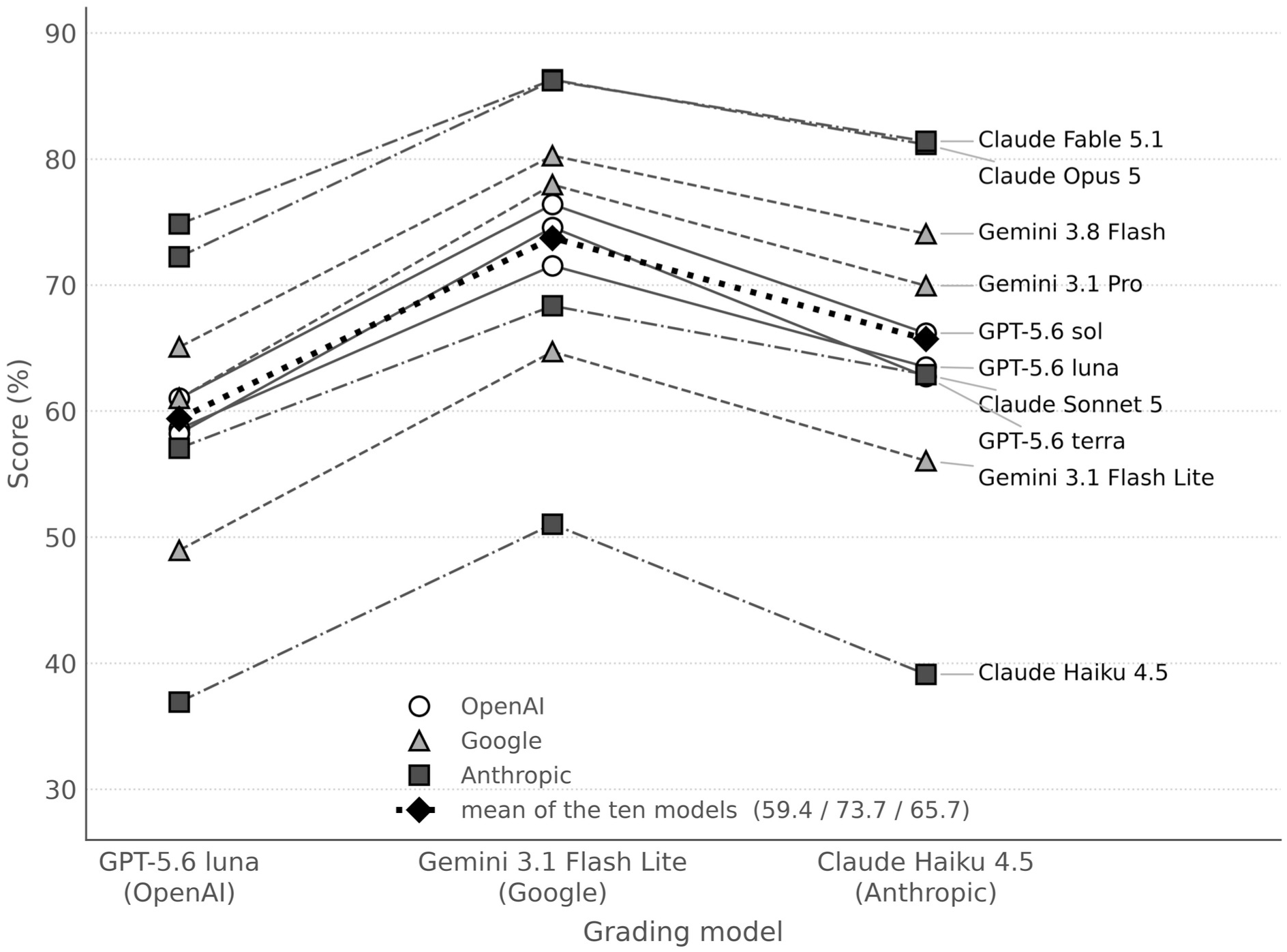
The same 740 responses, scored by three grading models. Each line is one of the ten evaluated models; the dotted line is their mean. The lines rise and fall together and rarely cross (Spearman’s rho 0.952-0.976 between graders, no model moving more than two places), so the ranking is a property of the responses. The level, however, moves by 14.3 points between graders, so an absolute score is a property of the grader as much as of the model. Alt text: Ten lines, one per evaluated model, connecting the scores given by the OpenAI, Google and Anthropic graders. Every line rises from the OpenAI grader to the Google grader and falls again at the Anthropic grader, and the lines keep their order; their mean moves from 59.4% to 73.7% to 65.7%.

### No family favoritism was detected

Each grader’s mean deviation from the models’ three-grader means was more favorable to its own family by 0.4 percentage points for the OpenAI grader, 0.6 for the Google grader and 0.4 for the Anthropic grader (Table 3). All three point the same way, but all three are smaller than the bootstrap standard deviation of a single measurement, and an order of magnitude smaller than the differences between graders in overall leniency. The highest-scoring models were Anthropic’s, and the Anthropic grader was not the most generous of the three.

**Table 3.**
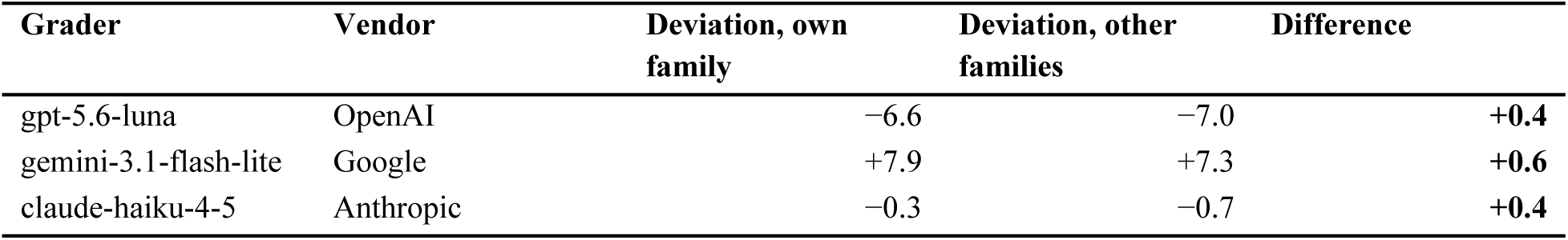
Test for grader family favoritism. For each model, the mean of its three grader scores was taken; the table gives each grader’s mean deviation from those means, separately for models of the grader’s own family and for models of the other two vendors. A positive difference indicates that the grader scored its own family more favorably. All three differences are smaller than the bootstrap standard deviation of a single measurement (2.6-4.1).

### How much a higher price buys differs sharply by vendor

From the cheapest tier to the most expensive, OpenAI’s models scored 58.6 → 58.2 → 61.0, Google’s 49.0 → 65.1 → 61.0 and Anthropic’s 36.9 → 57.1 → 74.8 (Figure 2). The cheapest OpenAI model beat the cheapest Anthropic model by 21.7 points, while the most expensive Anthropic model beat the most expensive OpenAI model by 13.8; Google’s mid-priced model outscored its own most expensive one. Which vendor looks best depends on the price tier at which the comparison is made.

**Figure 2.**
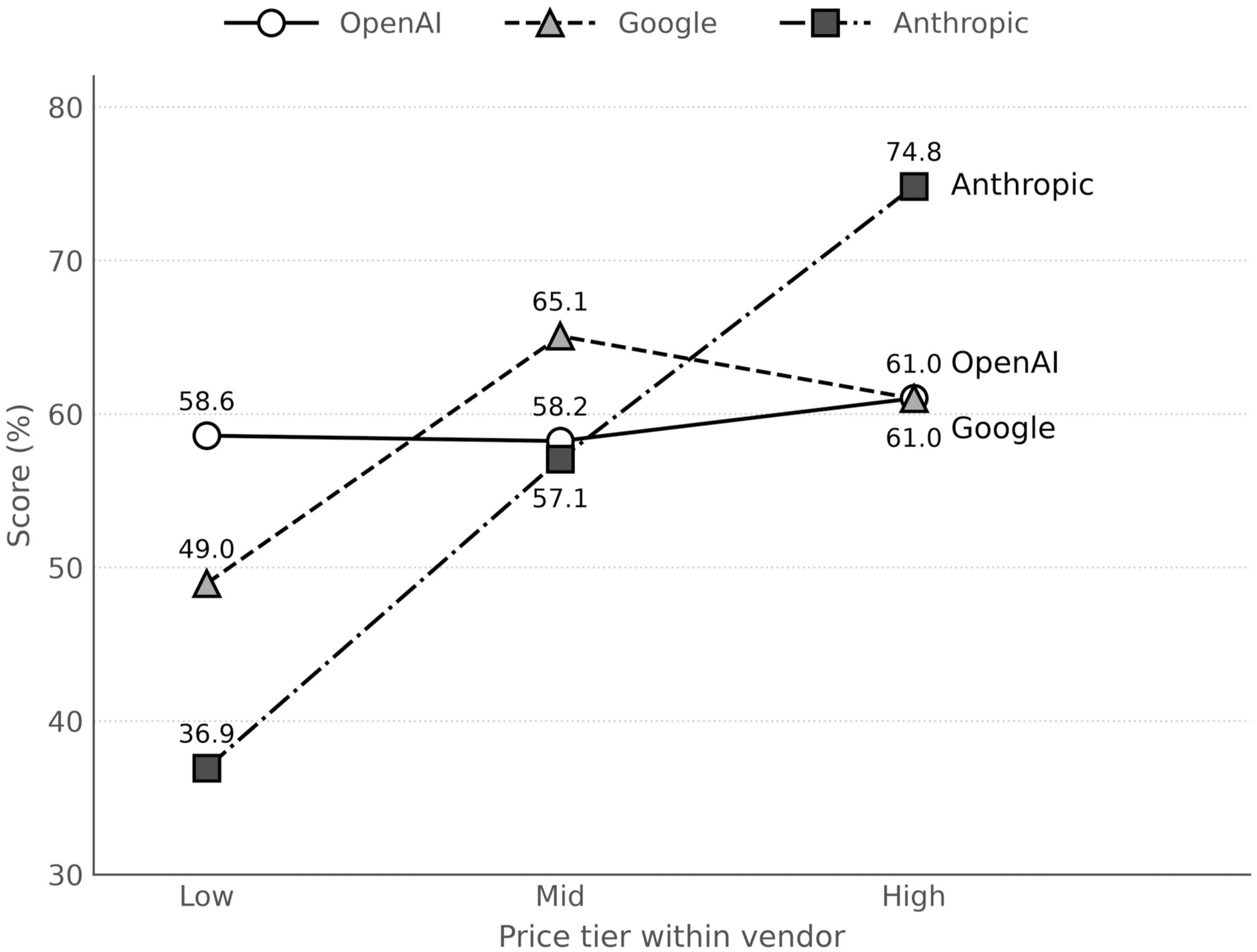
Score by price tier, within each vendor. Each vendor’s three models are plotted from its cheapest to its most expensive tier. Graded by GPT-5.6 luna. The trajectories differ in size and in shape: Anthropic’s models gain 37.9 points from the cheapest tier to the most expensive, OpenAI’s 2.4, and Google’s mid-priced model outscores its most expensive one. A comparison of vendors made at one price tier does not hold at another. Alt text: Line plot of benchmark score against price tier for three vendors. Anthropic rises steeply from 36.9% at the cheapest tier to 74.8% at the most expensive; OpenAI is almost flat, 58.6% to 61.0%; Google peaks at its mid tier, 65.1%, above its most expensive model at 61.0%.

### Where the models fail

Averaged over the ten models, themes ranged from emergency referrals at 83.6% down to response depth — a case described, a formula to be chosen and justified — at 41.9% (Figure 3a). By axis, accuracy was 64.3%, completeness 59.8% and context awareness 42.9%.

**Figure 3.**
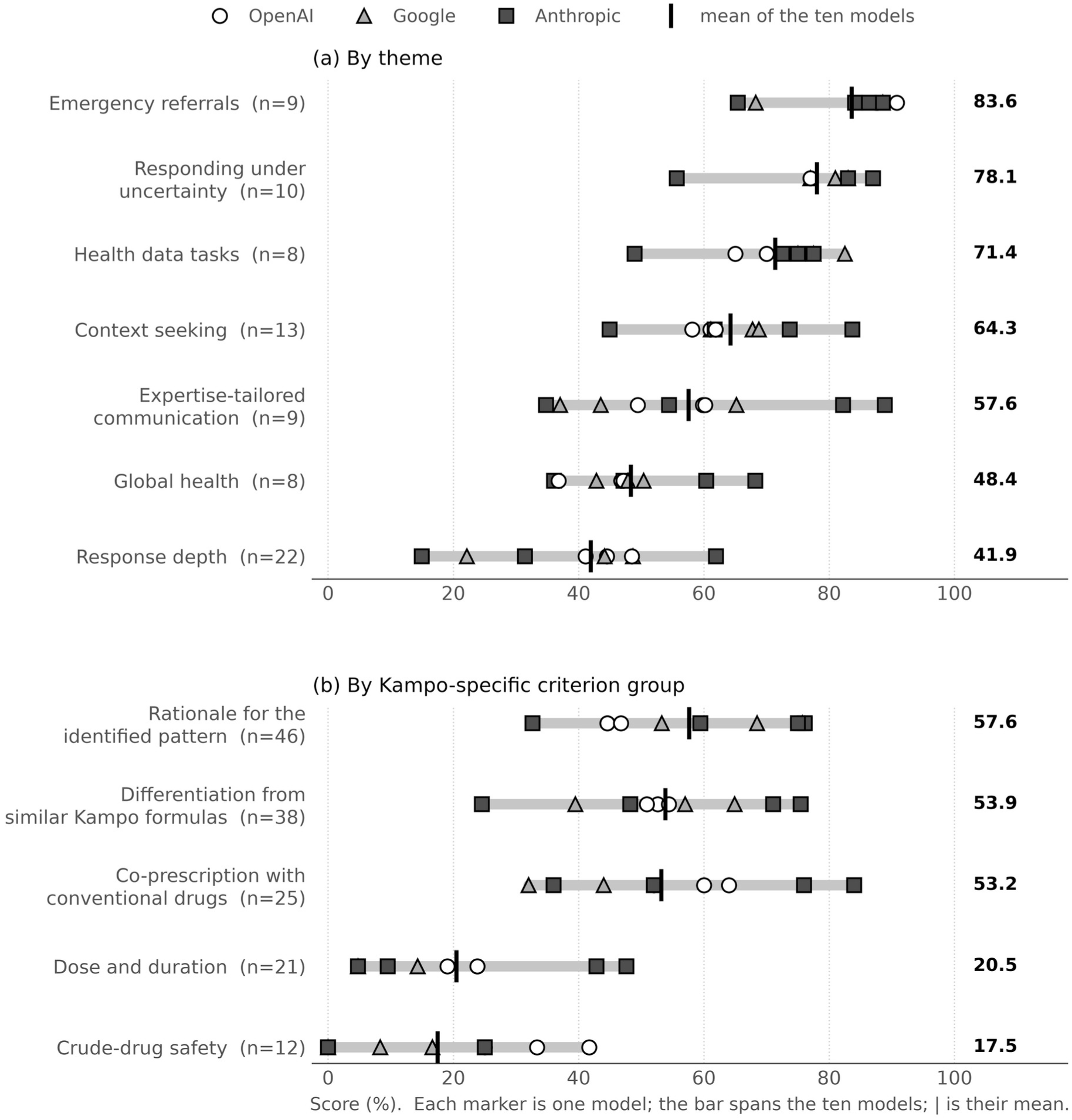
Where the models earn and lose points. Each marker is one of the ten models, the bar spans them, and the vertical rule is their mean, which is repeated at the right. Graded by GPT-5.6 luna; *n* is the number of scenarios in each group. (a) By theme: the models are most reliable when the required judgment is to advise that care be sought, and least reliable when a formula must be chosen and justified from a described case. (b) By Kampo-specific criterion group: dose and duration, and crude-drug safety, are markedly the weakest, and the ordering within them does not follow the overall ranking. Criteria in these two groups are compound (see Results 3.5). Alt text: Two panels of dots with a bar spanning them. Panel a gives the ten models’ scores for each of the seven themes, highest for emergency referrals at a mean of 83.6% and lowest for response depth at 41.9%. Panel b gives the five Kampo-specific criterion groups, highest for rationale for the identified pattern at 57.6% and lowest for crude-drug safety at 17.5%, where three models scored zero.

The Kampo-specific criteria separate more sharply (Figure 3b): rationale for the identified pattern 57.6%, differentiation from similar Kampo formulas 53.9%, co-prescription with conventional drugs 53.2%, dose and duration 20.5%, and crude-drug safety 17.5%, on which three models scored zero.

That last failure was not a failure of knowledge. Across the 120 gradings of the crude-drug-safety criteria, 95 responses (79.2%) named the crude drug at issue somewhere in the text, and 74 of those 95 still failed the criterion. A response was counted as naming the drug when the name of a crude drug the criterion concerns appeared anywhere in it: liquorice in ten of the twelve criteria, ephedra in one of those ten as well, scutellaria in one, and peach kernel or moutan bark in one. What the criteria ask is that the risk be tied to the case in front of the model — for example, whether any of the formulas the patient already takes contains that drug, what dose of it the patient would receive, and whether the amount taken needs to be monitored — and each asks for several such things at once. Criteria written in this compound form are common throughout the set, and depress absolute scores. These two groups are also small (12 and 21 scenarios), and the ordering within them did not follow the overall ranking: the highest score on crude-drug safety, 41.7%, belonged to gpt-5.6-luna, the cheapest OpenAI model, while claude-opus-5, the highest scoring model overall, scored 25.0%.

### Penalty criteria were almost never triggered

Of the 46 negative-point criteria, applied to ten models, exactly one was triggered — 1 of 460 — and none of the fifteen on the communication-quality axis. Current models do not make the errors those criteria were written to catch.

### The criteria are attainable

Scored against the same criteria, the physician-revised reference answers reached 87.9% (bootstrap SD 2.3), 92.4% (1.8) and 89.0% (2.2) under the three graders, with 49, 58 and 51 of the 74 scenarios at full marks. The ordering follows the graders’ leniency in Table 2, and the gap to the best model was 13.1, 6.1 and 7.6 points. The criteria are therefore attainable, and the models’ scores are not an artefact of criteria that no answer could satisfy. The reference answers were also shorter than the model responses (median 513 characters against 898-2,895). LLM graders can show verbosity bias, awarding higher scores to longer answers simply for their length [18]; here the shorter answers scored higher, so that bias is not what produced the result. The reference answers scored lower on context awareness than on the other axes (67.1%, 83.3% and 74.3%, against 88-95% for accuracy and completeness), as the models did. When even a physician-revised answer does not satisfy these criteria, what they ask is likely to be beyond what any single answer can show.

## Discussion

### What a consultation benchmark measures that an examination cannot

The criteria the models met most often were those about telling the user to seek care; the ones they met least often were those requiring a known risk to be tied to the case at hand. That is the gap an examination cannot see. The knowledge itself is not missing — the models named the crude drug in most of the responses that failed — but naming a risk is not the same as working out that two of the formulas in front of the patient contain it, at what daily dose, and what to watch for. Examination-format evaluations ask for the first, and models now answer them well [6,10]. A patient asking whether to add a second formula to what they already take is asking for the second.

### Grader family bias was not detected, but grader leniency is large

We built the three-grader design because a benchmark whose scores come from one vendor’s model invites the objection that it flatters that vendor, and because the effect is documented [16,17]. It was not detectable here: the difference between how a grader treated its own family and how it treated the others was well inside the noise of a single measurement. All three deviations point the same way, so we do not claim the effect is zero, only that it is too small here to change any conclusion.

Leniency is the larger problem. The same 740 responses averaged 59.4%, 73.7% and 65.7% depending on who graded them, and the reference answers reproduced the same ordering, which suggests that the graders differ in threshold rather than in what they respond to. The practical consequence is that a ranking can be had cheaply — our three graders are all low- or mid-priced models, self-preference is reported not to track grader capability [20], and mid-tier graders match frontier ones at much lower cost [21] — whereas an absolute number, of the kind a decision to deploy would need, requires the grader to be fixed and named, and the threshold argued for separately.

### Measure the tier you intend to pay for

The three vendors’ models behave differently as price rises, to the point that a comparison made at one tier reverses at another and one vendor’s mid-priced model beats its own flagship. Anyone selecting a model for a Kampo-related application should therefore measure the tier they intend to pay for. A benchmark that reports one model per vendor cannot answer the question they are actually asking.

### The distance to a physician-revised answer

The best model is 6 to 13 points short of the physician-revised answers, depending on the grader; the gap is real but not large, and it is smallest under the most lenient grader, which is a further reason to fix the grader before quoting a gap.

It is more informative to ask which criteria separate the two, and which defeat both, than to ask how large the overall gap is. Context awareness is weak for the reference answers as well as for the models, which points at the criteria on that axis rather than at the answers. On the compound safety criteria, by contrast, the reference answers do well and the models do not — and that is where the benchmark is carrying information.

### What the benchmark does not yet distinguish

One negative-point criterion was triggered in 460 opportunities. These criteria were written to catch responses that steer a patient away from medical care — urging them to stop a prescribed drug without medical advice, or to wait and see when they should be seen — responses that recommend a formula despite a contraindication, and responses that press for intimate detail beyond what a first consultation needs. Current models did none of these in response to our prompts. Fifteen of the sixteen communication-quality criteria are of this kind, which is why that axis cannot be reported as a percentage and contributes nothing to the ranking. A revision of the benchmark should either retire these criteria or write scenarios that put them under real pressure.

### Limitations

**Scale.** 74 scenarios and 310 criteria is a pilot. The subgroup results, and the Kampo-specific groups in particular at 12 to 46 scenarios, indicate directions to look rather than estimates.

**Single-turn.** 73 of the 74 scenarios are single-turn, so the benchmark does not measure whether a model asks a follow-up question and uses the answer. Given that context awareness was the weakest axis, this is the most important thing missing.

**Compound criteria.** Many criteria ask for several things at once and are graded as a single binary judgment, which depresses absolute scores and obscures which requirement was missed. In one scenario the physicians revised the reference answer without the corresponding criterion being updated, leaving the two contradicting each other — an inconsistency that a procedure of LLM drafting plus expert review can leave behind.

**Provenance of the material.** Scenarios, criteria and reference answers derive from drafts written by one model family, so an advantage to models of that family cannot be excluded, although its models did not lead the ranking under any grader. The reference answers are not a physician baseline: they were revised rather than written by physicians, those physicians are leading figures in the field rather than a representative sample, and they also reviewed the criteria.

**Grading was not validated against physicians.** Agreement between three graders of different families detects disagreement, but not error they share.

## Conclusion

We built KampoBench, a Japanese benchmark of 74 consultation scenarios with 310 physician-reviewed criteria, and measured ten current large language models with three graders drawn from three different vendors. Rankings were stable across graders and no family favoritism was detectable, but grader leniency varied by 14.3 points, so a score is meaningful only alongside the grader that produced it. The best model reached 74.8-86.3%, against 87.9-92.4% for the physician-revised reference answers. The models were most reliable at advising that care be sought and least reliable at tying a known risk — crude-drug safety, dose and duration — to the case in front of them, a distinction that examination-format evaluations cannot make. That distinction is what a benchmark for traditional medicine now needs to measure.

## Author contributions

Following the CRediT taxonomy. H.T.: conceptualization, methodology, software, formal analysis, data curation, investigation, visualization, writing - original draft, project administration. Y.H., H.I., K.U. and T.Y.: validation, data curation, writing - review and editing. T.N.: validation, data curation, resources, supervision, writing - review and editing. All authors read and approved the final manuscript.

## Acknowledgements

We thank the External Affairs Committee, The Japan Society for Oriental Medicine, for its advice and for making board-certified Kampo physicians available for this work.

The authors used a large language model (Claude, Anthropic) to draft and edit portions of this manuscript. All content was reviewed and verified by the authors, who take full responsibility for it. The use of a large language model to draft the dataset itself is described in Materials and Methods.

## Ethics

This study involved no human participants and no patient data; the scenarios are synthetic. Ethical approval was therefore not required.

## Data availability

The dataset (74 scenarios, 310 criteria) is deposited at Zenodo under CC BY 4.0, doi:10.5281/zenodo.23129442 [22]; the files are restricted until the article is published in a journal and will be opened then. Every example carries a canary string, embedded in September 2026; please do not repost the contents as plain text or as images, so that the benchmark is not absorbed into training corpora. The same record contains the per-criterion grading outcomes for all ten models and the reference answers under the three graders, from which the tables and figures can be reproduced. The model responses and the graders’ explanations are not deposited, to keep the benchmark out of training corpora; they are available from the corresponding author on request. The evaluation code, the measurement conditions and the provenance of the dataset are at https://github.com/medicalcloud/kampobench.

## References

1. Ministry of Internal Affairs and Communications. Outline of the 2026 White Paper on Information and Communications in Japan. Tokyo: MIC; 2026. https://www.soumu.go.jp/johotsusintokei/whitepaper/eng/WP2026_overview.pdf

2. Japan Cancer Society. Survey Report on the Use of Generative AI by Cancer Patients and Their Families. Tokyo: Japan Cancer Society; 2026. https://www.jcancer.jp/gsclub/research_260618 (in Japanese)

3. Cai X, Li W, Shi W, et al. Behavioral dynamics of AI trust and health care delays among adults: integrated cross-sectional survey and agent-based modeling study. J Med Internet Res. 2026;28:e82170. doi:10.2196/82170

4. KFF. KFF Tracking Poll on Health Information and Trust: Use of AI for Health Information and Advice. San Francisco, CA: KFF; 2026. https://www.kff.org/public-opinion/kff-tracking-poll-on-health-information-and-trust-use-of-ai-for-health-information-and-advice/

5. Moschik EC, Mercado C, Yoshino T, et al. Usage and attitudes of physicians in Japan concerning traditional Japanese medicine (Kampo medicine): a descriptive evaluation of a representative questionnaire-based survey. Evid Based Complement Alternat Med. 2012;2012:139818. doi:10.1155/2012/139818

6. Yue W, Wang X, Zhu W, et al. TCMBench: a comprehensive benchmark for evaluating large language models in traditional Chinese medicine. arXiv. 2024. arXiv:2406.01126, preprint: not peer reviewed.

7. Huang T, Chen J, Lu L, et al. TCM-5CEval: extended deep evaluation benchmark for LLM’s comprehensive clinical research competence in traditional Chinese medicine. arXiv. 2025. arXiv:2511.13169, preprint: not peer reviewed.

8. Iizuka N, Usuku K, Nakae H, et al. Web-based evaluation system to measure learning effectiveness in Kampo medicine. Evid Based Complement Alternat Med. 2016;2016:2043535. doi:10.1155/2016/2043535

9. Iizuka N, Nakae H, Segawa M, et al. Web-based evaluation system for closing the education gap in Kampo medicine between facilities. Tradit Kampo Med. 2019;6(1):12–18. doi:10.1002/tkm2.1207

10. Takagi S, Watari T, Erabi A, et al. Performance of GPT-3.5 and GPT-4 on the Japanese Medical Licensing Examination: comparison study. JMIR Med Educ. 2023;9:e48002. doi:10.2196/48002

11. Shimada Y, Fujimoto M, Nogami T, et al. Adverse events associated with ethical Kampo formulations: analysis of the domestic adverse-event data reports of the Ministry of Health, Labor, and Welfare in Japan. Evid Based Complement Alternat Med. 2019;2019:1643804. doi:10.1155/2019/1643804

12. Shimada Y. Adverse effects of Kampo medicines. Intern Med. 2022;61(1):29–35. doi:10.2169/internalmedicine.6292-20

13. Wei J, Huang D, Lu Y, et al. Simple synthetic data reduces sycophancy in large language models. arXiv. 2023. arXiv:2308.03958, preprint: not peer reviewed.

14. Arora RK, Wei J, Soskin Hicks R, et al. HealthBench: evaluating large language models towards improved human health. arXiv. 2025. arXiv:2505.08775, preprint: not peer reviewed.

15. Takata T, Yamada R, Rene AON, et al. Development of a virtual patient model for Kampo medical interview: new approach for enhancing empathy and understanding of Kampo medicine pathological concepts. In: 2024 Joint 13th International Conference on Soft Computing and Intelligent Systems and 25th International Symposium on Advanced Intelligent Systems (SCIS&ISIS). Himeji, Japan: IEEE; 2024:1–5. doi:10.1109/SCISISIS61014.2024.10759962

16. Panickssery A, Bowman SR, Feng S. LLM evaluators recognize and favor their own generations. arXiv. 2024. arXiv:2404.13076, preprint: not peer reviewed.

17. Spiliopoulou E, Fogliato R, Burnsky H, et al. Play favorites: a statistical method to measure self-bias in LLM-as-a-judge. arXiv. 2025. arXiv:2508.06709, preprint: not peer reviewed.

18. Zheng L, Chiang WL, Sheng Y, et al. Judging LLM-as-a-judge with MT-Bench and Chatbot Arena. In: Advances in Neural Information Processing Systems 36 (NeurIPS 2023) Datasets and Benchmarks Track; 2023. arXiv:2306.05685

19. Chen GH, Chen S, Liu Z, et al. Humans or LLMs as the judge? A study on judgement biases. arXiv. 2024. arXiv:2402.10669, preprint: not peer reviewed.

20. Yang J, Hu Z, Qiu C, et al. Quantifying and mitigating self-preference bias of LLM judges. arXiv. 2026. arXiv:2604.22891, preprint: not peer reviewed.

21. Soumik SK. Judging the judges: a systematic evaluation of bias mitigation strategies in LLM-as-a-judge pipelines. arXiv. 2026. arXiv:2604.23178, preprint: not peer reviewed.

22. Takata H, Harada Y, Inoue H, Uneda K, Yoshino T, Nogami T. KampoBench: a physician-reviewed Japanese benchmark for large language models in Kampo medicine [data set]. Zenodo; 2026. doi:10.5281/zenodo.23129442

